# PCR Testing in the UK During the SARS-CoV-2 Pandemic – Evidence from FOI Requests

**DOI:** 10.1101/2022.04.28.22274341

**Authors:** Tom Jefferson, Madeleine Dietrich, Jon Brassey, Carl Heneghan

## Abstract

Polymerase Chain Reaction (“PCR”) tests have been used to identify cases of COVID-19 during the course of the pandemic. Notably, PCR alone cannot differentiate between the presence of whole viruses (which can be transmitted and infect individuals) and small fragments of genetic material that are not infectious. A feature of PCR known as the cycle threshold (Ct) can be used to discriminate between these states, but the relationship between Ct and infectiousness is still poorly understood.

This well-known limitation of the test compromises the identification of cases and their trends, and consequently those measures to interrupt transmission (such as isolation) that are undertaken on the basis of reliably identifying infectious individuals.

Here, we interrogate the public authorities’ understanding of PCR testing for SARS-CoV-2 in the UK by accessing Freedom of Information requests submitted in 2020-21.We searched <u>WhatDoTheyKnow</u> and found <u>300 FOI requests</u>, from over 150 individuals. We grouped their questions into four themes addressing the number of tests in use, the reporting of cycle thresholds (‘Ct’), the Ct values themselves, and the accuracy of tests.

The number of <u>validated</u> tests in use in the UK is currently not clear: In FOI responses, Public Health England (PHE) report it may be “<u>80</u>” or “<u>85</u>”. However, European regulations suggest there could be <u>over 400 different CE marked tests available on the market</u> and available for use. Laboratories have a statutory duty to report positive cases to PHE, but they do not have to advise which tests they are using nor submit Ct values. Only two FOI responses provided answers on Ct values, indicating that in a set time span, 24–38% of the Ct values were over 30. The most common FOI asked if there was a cycle threshold for positivity. In those that responded, the Ct for a positive result varied from 30 to 45. We found limited information on the technical accuracy of the tests. Several responses stated there is no ‘static’, ‘specific’ or ‘standard’ cycle threshold.

The current system requires significant changes to ensure it offers accurate diagnostic data to enable effective clinical management of SARS-CoV-2. PCR is an important and powerful tool, but its systematic misuse and misreporting risk undermining its usefulness and credibility.

## Introduction

PCR (polymerase chain reaction) testing has played a central role in tracking the progress of the SARS-CoV-2 pandemic, and in enabling the restriction measures that were put in place to suppress the spread.

PCR is a technique that “amplifies” small segments of genetic material to identify an infectious agent. The results of a PCR test are qualitative: either positive or negative. The result is based on whether the targeted genetic material is identified and how many amplification cycles are required to identify it. The cycle threshold (Ct) of the PCR test is the number of amplification cycles at which the sample yields a positive result.

The smaller the quantity of (targeted) genetic material present in the test sample, the harder the PCR must work to find it, i.e., the more cycles of amplification it has to perform. The technique becomes more sensitive as the number of amplification cycles is increased –until it’s searching for a needle in a haystack. The Ct is set by the laboratory or test programme, and as Ct is raised, the likelihood of a positive test result will increase for a given sample.

The UK Health Security Agency <u>guidance</u> reports, “samples with Ct<25 are considered to have a high amount of virus, those with Ct>25<30 are considered to have a medium amount of virus, and those with Ct>30 are considered to have a low amount of virus.”‘ Therefore, when the Ct is set at 45, the test can detect single gene fragments and render these as a positive result even though gene fragments themselves are not infectious and can linger in the patient long after the infection has passed.

However, PCR alone cannot differentiate between the presence of whole viruses (which can be transmitted and infect individuals) and small fragments of genetic material that are not infectious. The shedding of this viral debris can occur over a prolonged period of time.

To identify infectious individuals, the interpretation of the cycle threshold should be undertaken in the context of symptom onset and severity, age, and past medical history. Broadly, an individual is likely to be infectious with a Ct below 30.^1^ Furthermore, a week or more after the onset of symptoms, the average person is no longer likely to be infectious. Those with a weakened immune system are likely to be infectious for longer.

The central importance of PCR, the cycle threshold, and its relationship to infectiousness and, thus, containment measures have not escaped notice by several members of the public, and this has resulted in a substantial number of Freedom of Information Act (“FOIA”) requests for data to substantiate the PCR results.

The Freedom of Information Act of 2000 allows for a public “right of access” to information held by public authorities in the UK. A request can be submitted by contacting a public body such as a government department or hospital trust directly or by using the website <u>WhatDoTheyKnow</u>, which forwards requests to the appropriate authority and publishes the subsequent responses.

We, therefore, collated FOI responses posted on the <u>WhatDoTheyKnow</u> site in 2020 and 2021 to understand the mechanisms by which PCR testing is governed and how the agencies delivering testing understand their use and are accountable to the public.

## Methods

We set out to analyse public authorities’ understanding of PCR testing for SARS-CoV-2 in the UK by accessing Freedom of Information requests posed in 2020-21. By public authorities, we mean NHS Health Trusts, laboratories, and government agencies such as Public Health England and the Department of Health and Social Care.

We searched <u>WhatDoTheyKnow</u> using the terms “PCR cycle threshold” to understand the use of cycle thresholds in the UK public sector and found <u>300 FOI requests</u>, from over 150 individuals.

We grouped questions into four themes addressing the number of tests in use, the reporting of cycle thresholds, the cycle threshold values, and the accuracy of tests.

These were in our view the most important issues. However, the richness of the questions and answers can be fully appreciated <u>in our extraction</u>.

| Themes & Questions |  |
| --- | --- |
| 1. | Test Numbers in Use: |
| 1.1 | **How many different PCR tests kits are being used in the UK?** |
| 2. | Cycle Threshold Reporting: |
| 2.1 | **Is there statutory reporting of Ct values with the results of the PCR test?** |
| 2.2 | **Is the PCR cycle threshold required to be recorded with every PCR test?** |
| 3. | Cycle Threshold Values: |
| 3.1 | **Do you know what the Ct values are?** |
| 3.2 | **Do you have a Ct threshold for positivity?** |
| 4. | PCR Test Accuracy: |
| 4.1 | **Can you approximate the false positive and false negative rates for your PCR test? And does this vary with the Ct?** |

We analysed all the retrieved FOIA requests by extracting data relevant to the questions; to whom they were directed with the date of the request, and the response along with the answers, and we selected relevant quotes from the answers. We categorised responses into six domains: ‘Cycle Threshold (“Ct”)’, ‘Ct Change’, ‘PCR - Other information’, ‘Case Definition’, ‘C19 Deaths’, and ‘Other’. One author (TJ, MD or CH) independently extracted data into a google sheet and the data was checked by a second reviewer.

### Data Sharing

All the FOI responses are accessible <u>here</u>.

## Results

### 1. Test Numbers in Use

#### 1.1 How many different PCR test kits are being used in the UK?

None of the FOIA requestors asked directly for the type or number of tests used in the UK. However, in responses to questions about cycle thresholds and general PCR information, eleven unsolicited answers were provided.

##### Hospital Trusts

Seven answers were from NHS Trusts: six respondents provided the names of the PCR tests, and one response from Belfast Health and Social Care Trust reported that numerous PCR based assays were used (See Table 1.1).

**Table 1.1:** NHS Trusts responses to how many tests are being used.

| Responder and Response | Link |
| --- | --- |
| **Belfast Health and Social Care Trust:** The numerous PCR based assays for SARS-CoV2 used by the Regional Virus Laboratory in Belfast Trust employ Taqman technology. | Link |
| **North Cumbria Integrated Care NHS Foundation Trust:** 20.03.2020 Seegene Seeprep12/Applied biosystems ABI 7500.; 08.04.2020 Roche Cobas 6800. 04.06.2020 Seegene STARlet. Number of thermocycles =44; 17.12.2020 Seegene STARlet. Number of thermocycles = 2 initial thermocycles, 41 thereafter ** Ct Change** 30.11.2020 Altona AltoStar; 05.05.2020 Cepheid geneXpert. | Link |
| **Guy's and St Thomas' NHS Foundation Trust:** Two rapid PCR assays are in use within the Viapath laboratories, the Cepheid Xpert SARS-CoV-2 assay and the MobiDiag NovoDiag COVID-19. These are “black box” analysers, and users are not provided with full details of assay design and number of cycles used. | Link |
| **Leeds Teaching Hospitals NHS Trust:** Two kits are in use at the time of the request: NeuMoDx SARS CoV-2 in November 2020: Cepheid Gene Xpert in use June 2020, Cts respectively up to 30 and 35. | Link |
| **Countess of Chester Hospital NHS Foundation Trust:** Samba II - Doesn't record cycles; Primer Design - 40 cycles; Cepheid - 45 cycles; Respibio - 45 cycles; Biofire - Doesn't record cycles | Link |
| **Greater Glasgow NHS Board:** Amplification cycles in PCR test for COVID Glasgow Royal Infirmary Applied Biosystems; 7500 Real-Time PCR System 40; Abbott Alinity M 40; Roche Cobas 6800 45; Seegene CFX96 45; Cepheid GeneXpert 45; Abbott M2000 40; Roche Liat 45; QEUH Roche Liat 45; Hologic; Panther n/a**; Thermo Fisher Scientific Amplitude 40. | Link |
| **Blackpool Teaching Hospitals NHS Foundation Trust:** Manufacturer/assay and remains constant, as we cannot amend the software, however confirmation of positive results is performed where the sample Ct threshold is >38 on Cepheid Genexpert and >34 Neumodx analysers. | Link |

##### Public Health England Responses

Five further responses were provided by Public Health England (see Table 1.2). From January through to August 2021, PHE responded that at least 80 different platforms were in use, configured in various ways in different laboratories. By September this number had increased to 85.

**Table 1.2:** PHE Assessment & CE Mark Status for Tests Reported by Trusts per the February 2021 PHE Report.

| Name of Test | PHE Assessed | CE Marked |
| --- | --- | --- |
| Abbott Alinity M | No | Yes |
| Abbott M2000 | No | No |
| Altona AltoStar. | Yes | Yes |
| Applied Biosystems 7500 Real-Time PCR System | No | Yes |
| Cepheid geneXpert | No | Yes |
| Hologic; Panther | No | No |
| MobiDiag NovoDiag COVID-19 | No | Yes |
| NeuMoDx SARS CoV-2 | No | Yes |
| PrimerDesign (Novacyt) | Yes | Yes |
| QEUH Roche Liat | No | No |
| Respibio | No | No |
| Roche Cobas 6800 | Yes | Yes |
| Roche Liat | No | No |
| Seegene CFX96 | No | No |
| Seegene Seeprep12 | Yes | Yes |
| Seegene STARlet. | No | No |
| Thermo Fisher Scientific Amplitude | No | No |

However, of the 80 or 85 tests, <u>17</u> had been reported as assessed by PHE by February 2021. Of the 17 tests reported by the seven trusts (Table 1.1), four were PHE assessed and nine were CE marked. We were able to verify that four tests were both PHE assessed and CE marked: Altona AltoStar, PrimerDesign (Novacyt), Roche Cobas 6800, and Seegeene Seeprep 12. Eight tests were neither PHE assessed or CE marked (see Table 1.2 and text below).

Further <u>guidance</u> from the ‘National technical validation process for manufacturers of SARS-CoV-2 (COVID-19) tests’ (updated 2 February 2022) reports 24 ‘validated’ technologies in the pipeline to support the test and trace program. This <u>report</u> on the findings from technical validations and in-service valuations, last updated in December 2021, cites 22 test ‘validations’.

In a December 2020 response, <u>PHE confirmed they held information</u> on testing kits used by PHE laboratories. In the same response, PHE confirmed they do *not* hold information on testing kits used by non-PHE laboratories, despite these laboratories having a statutory duty to report positive cases to PHE.

The UK government information on <u>national technical validation</u> reports that

> ‘*whether or not a test developer or supplier explores national-level procurement via this process, they are still able to supply tests with the relevant regulatory authorisation to UK customers*.’

In the UK, all medical devices, including in vitro diagnostic medical devices (IVDs), <u>must be registered with the MHRA before they are placed on the market</u>. CE marking will continue to be recognised in Great Britain until 30 June 2023.

In the 360dx <u>Coronavirus Test Tracker</u> of Commercially Available COVID-19 Diagnostic Tests, there are currently 271 <u>with a CE mark</u> of which 141 are PCR tests that can be legally sold and made available in the UK.

However, according to the <u>European Commission’s</u> COVID-19 In Vitro Diagnostic Devices and Test Methods Database, there are 2,136 CE marked diagnostic tests, 420 of which are PCR tests.

An additional list - published by the Department of Health & Social Care and the UK Health Security Agency, also published on 18 October 2021 (updated on 3 November) - includes <u>48 tests which are exempt from validation requirements</u> until 28 February 2022.

The number of ‘validated’ tests is therefore not clear: it seems that PHE assessment ceased in January 2021. The repeatedly used standard response (boilerplate) of “80” such tests in use started in the same month. However, we are unclear where this number came from as there is no list of the tests, or the names of the additional tests that gave rise to the increase to 85.

The DHSC published a COVID-19 test approval and <u>how to apply</u> on 28 July 2021. Within this approval process, the Medical Devices (Coronavirus Test Device Approvals) 2021 <u>Regulations</u> set out that ‘*manufacturers or distributors supplying COVID-19 tests must apply to the Department of Health and Social Care (DHSC) for approval*.’ The DHSC review assesses the evidence a supplier submits against a minimum required data set - known as a ‘desktop review’. DHSC only <u>publishes details of tests that have passed</u>.

The list includes 16 tests (see Table 1.3). These tests were approved in accordance with <u>regulation 38A</u> between 14 Oct 2021 and 10 January 2022.

**Table 1.3.** DHSC: <u>COVID-19 test validation approved products</u> as of 10 Jan 2022.

|  |
| --- |
| Bosphore Novel Coronavirus (2019-nCoV) Detection Kit v4 |
| Bosphore Novel Coronavirus Detection Kit v2 |
| Bosphore SARS-CoV-2- Respiratory Pathogens Panel kit v1 |
| Bosphore SARS-CoV-2-Flu-RSV Panel Kit |
| Bosphore SARS-CoV-2-Flu-RSV Panel Kit v2 |
| Cobas SARS-COV-2 + Flu A + Flu B for use with the cobas 6800/8800 Systems |
| Cobas SARS-COV-2 for use with cobas 6800/8800 systems |
| Coronavirus COVID-19 genesig Real-Time PCR assay |
| FluoroType®SARS-CoV-2/Flu/RSVVER 1.0 |
| Logix Smart SARS-CoV-2 (genes RdRp/E) |
| RealStar SARS-CoV-2 RT-PCR Kit 1.0 |
| RIDAGENE Flu A/B & SARS CoV-2 |
| TaqPath(TM) COVID-19 CE-IVD RT-PCR Kit |
| TaqPath™ COVID-19, FluA/B, RSV Combo Kit |
| VIASURE SARS-CoV-2 Real-Time PCR Detection Kit |
| Vivalytic SARS-CoV-2 |

**Table 1.4:** PHE responses to the question of how many tests are being used.

| Date of Response | Response | Link |
| --- | --- | --- |
| 4 Jan 2021 | ‘There are multiple tests being used and every kit will have its own interpretative criteria and thresholds, as recommended by the manufacturer. There are at least 80 different platforms in use, configured in various ways in different laboratories.’ | Link |
| 16 Feb 2021 | ‘Currently, there are over 80 different testing platforms in use across the various networks of laboratories and the thresholds for result interpretation will be configured in each case by the manufacturers of the assays and the instrumentation.’ | Link |
| 30 Jun 2021 | ‘There are over 80 different PCR tests and several platforms used in the UK – each comes with its own standardised interpretation – some provide Ct values, some not.’ | Link |
| 12 Aug 2021 | ‘There are multiple tests being used, and every kit will have its own interpretative criteria and thresholds, as recommended by the manufacturer. There are at least 80 different platforms in use, configured in various ways in different laboratories. Some test provides Ct values; some does not.’ | Link |
| 13 Sep 2021 | ‘There are over 85 different SARS-CoV-2 RT-PCR assays/platforms in use across the UK. Most tests used in England are commercial. Each test/platform comes with its own standardised interpretation and has a slightly different limit of detection.’ | Link |

The list of <u>European Commission’s</u> COVID-19 In Vitro Diagnostic Devices suggest there could be over 400 COVID-19 Diagnostic tests available on the market and available for use.

Table 1.2 shows that Trusts list non-PHE assessed tests, some of which are CE marked but also several that are not. We found no data to clarify if these tests are in the development phase or awaiting classification. Five of the 17 were, however, included on the exempted list.

We were unable to find a list of technical performance data for the 85 tests PHE refers to or whether these tests are currently CE marked, or indeed which tests are in use, and which have been abandoned.

PHE Technical Briefings <u>report</u> the Thermopath TaqPath assay is used in three <u>UK lighthouse laboratories</u>. According to the <u>UKHSA</u>, these mega throughput labs account for one-third of COVID PCR testing from July to Nov 2021 in England.

### 2. Cycle Threshold Reporting

#### 2.1 Is there statutory reporting of Ct values with the results of the PCR test?

Public Health England responded to fifteen FOIA requests relating to the reporting of Ct Values (see examples in Box 2.1). Their response indicates that while PHE and non-PHE laboratories have a statutory duty to report positive cases to PHE, they do not have to report which tests they are using nor submit Ct values to PHE.

##### Box 2.1

Sample requests & responses

<u>FOI request by Vaughan on 8 Jun 2021</u>:

> ‘W*hich of the following variables are recorded by PHE: test brand, the assay used, cycle threshold or clinical history*?’

<u>PHE response 6 July 2021</u>:

> *‘PHE does not hold information on the SARS-CoV-2 RT-PCR test kits used by non-PHE laboratories. These laboratories have a statutory duty to report positive cases to PHE, but they are not obliged to advise PHE of which tests are being utilised nor submit cycle threshold values. ‘*

*\*\*\**

<u>FOI request by Anderson 17 April 2021</u>

> *requested details on the minimum cycle threshold, the maximum cycle threshold and the average (mean) cycle threshold used in the PCR test to determine covid cases in the U*.*K*.

<u>PHE response 18 May 2021</u>:

> *‘These laboratories have a statutory duty to report positive cases to PHE, but they are not obliged to advise PHE which tests they are using nor submit CT values used to PHE*.*’*

#### 2.2 Is the PCR cycle threshold required to be recorded with every PCR test?

An FOI request <u>on 7 Dec 2020</u> solicited information on whether the PCR Ct is required to be recorded with every PCR test in England.

The Department of Health and Social Care (DHSC) <u>responded on 8 Jan 2021</u>:

> ‘*Ct values are not provided for all SARS-CoV-2 molecular detection methods. Some commercial RT-PCR techniques are closed ‘black box’ systems whereby the operator cannot observe the reaction in real-time and the result is interpreted by software into a qualitative non-interrogatable positive or negative result’*

An FOI to the Scottish Government <u>on the 14 Sep 2020 requested information</u> on the association of a positive test with a Ct. The response on 6 November stated:

> *‘The answer to your question is that Ministers haven’t directed labs on associating Ct value with a positive test, so we do not have a date to provide you with*.*’*
>
> *‘It is up to the virologists at the diagnostic labs to interpret the result of the test that is reported, and where a test looks to be weak positive then there are protocols in place to retest*.

The DHSC did not <u>respond on 17 September 2020</u> to a request as to whether the cycle threshold of COVID PCR tests is recorded for pillar 2 testing.

In February 2021, PHE <u>stated</u>

> “*Whilst each laboratory has a statutory duty to report positive cases to Public Health England, additional information such as Ct values in the case of PCR tests, is not submitted” and further* <u>added</u> *“nor are they obliged to advise what tests they are using*.’

### 3. Cycle Threshold Values

#### 3.1 Do you know what the Ct values are?

> ‘*The cycle threshold (Ct) can be defined as the thermal cycle number at which the fluorescent signal exceeds that of the background and thus passes the threshold for positivity*.’ see <u>Public Health England</u>

Fourteen FOIA requests asked for information on the Ct values of the PCR tests carried out by trusts and public health officials. Two respondents provided answers: in Wales in the month of October in 2020, 24% of the values were Ct >30 and in the Southern Health Care Trust (dates unclear) 38% > than a Ct of 30.

**Table 3.1:**
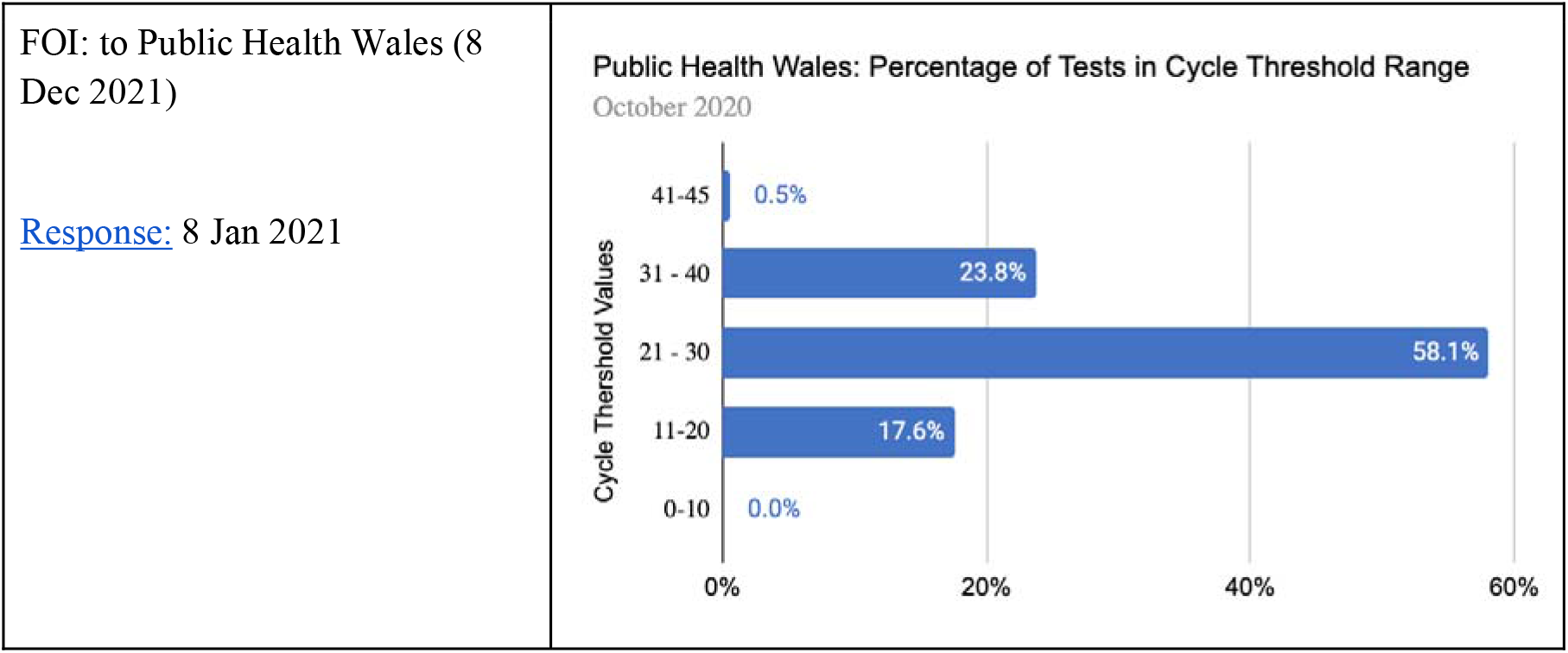
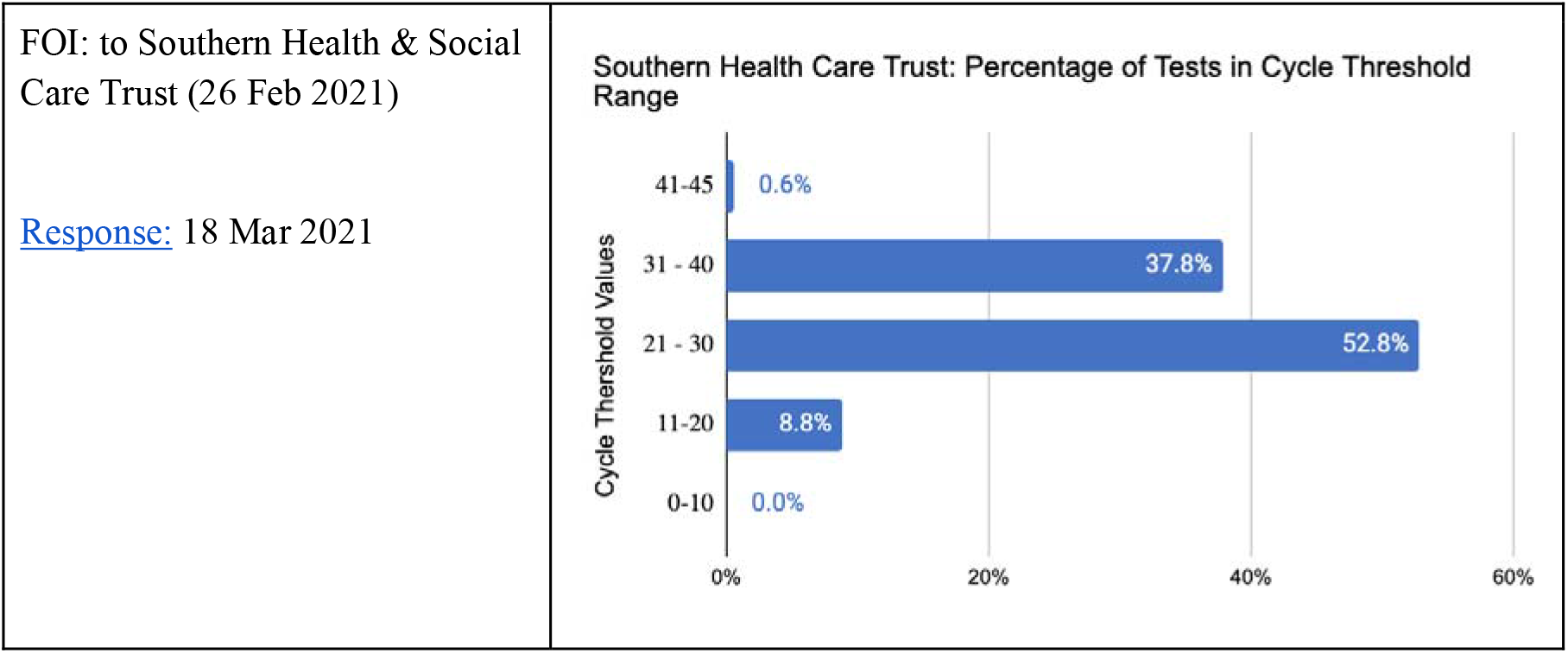
Percentage of Tests in the Cycle Threshold Range for Wales & Southern Healthcare Trust, per FOI response.

Non <u>responses</u> to FOI requests occurred due to an inability ‘*to produce the data due to the number of hours it would take for data systems to generate*.’

#### 3.2 Do you have a Ct threshold for positivity?

The request for a Ct threshold for positivity was the most common FOI with 184 requests. In the responses, the Ct considered a positive test varied from 30 through to 45 (see Table 3.2.1).

**Table 3.2.1.** Ct considered as a positive test.

| Response | Respondent |
| --- | --- |
| ‘A typical RT-PCR assay will have a maximum of **40 thermal cycles.**’ | DHSC |
| ‘As per manufacturer instructions, PCR assays are run as **40-45 cycles**, and the Ct cut off for determining a positive versus negative result is determined individually for every different SARS-CoV-2 assay run in laboratories. In very general terms, most will have a Ct cut-off of between **Ct 35-40** depending on the data and the assay.’ | Coventry & Warwickshire NHS Trust |
| ‘**45 Ct** used as standard in 2020 and 2021.’ | Mid and South Essex NHS Foundation Trust |
| ‘As stated previously, samples crossing the threshold at <**CT 37** are reported as positive and between >=CT 37 to <CT 40 results are reported as indeterminate and a repeat sample requested.’ | Belfast Health and Social Care Trust (Northern Ireland) |
| ‘We can confirm that our cycle threshold used with PCR tests is **30 – 35**’ | York and Scarborough Teaching Hospital NHS Foundation Trust |
| ‘The number of amplification cycles can vary with different platforms used. Most platforms use threshold cycles that range from **27\* to 43**. The threshold cycle is determined by the platform used and is not something that the laboratory services has control over.’ | Public Health Wales |
| ‘Of the primary assays that report a CT value, we report a positive result <38 CT. This is set by the manufacturer.’ | Hampshire Hospitals NHS Foundation Trust |
| ‘Range reported for different named tests **40-45**’ | Greater Glasgow NHS Board |
| 'The RT-PCR assays are run over **40 cycles**, and results are expressed as Cycle thresholds (CTs) as per the commercial manufacturer guidance.' | University Hospitals Sussex NHS Foundation Trust |
| 'The cycle threshold is **30-35**' | York Teaching Hospital NHS Foundation Trust |
| 'In cases where a PCR test is used, this runs at **35** thermal cycles as per the commercial manufacturer guidance.' | University Hospitals Sussex NHS Foundation Trust |
| 'Two kits in use at the time of the request: NeuMoDx SARS CoV-2 in use November 2020 Cepheid Gene Xpert in use June 2020, Cts respectively up to **30 and 35**' | Leeds Teaching Hospitals NHS Trust |
| '**40** cycles is routine' | Torbay and South Devon NHS Foundation Trust |
| 'three named tests use CT **40-45** and two tests, Samba II and Biofire. don't record cycles' | Countess of Chester Hospital NHS Foundation Trust |
| 'Each test runs **45** cycles as standard.' | Royal United Hospitals Bath NHS Foundation Trust |
| 'Western Sussex Hospitals NHS Foundation Trust threshold of a **CT of 34 is deemed detected**. Brighton and Sussex University Hospitals NHS Trust = PCR assays are run over **40 cycles**.' | University Hospitals Sussex NHS Foundation Trust |
| 'Typically a real-time PCR test will run for a maximum of around 40 thermal cycles - named tests have CT of **41-45**' | North Cumbria Integrated Care NHS Foundation Trust |
| 'The Ct value that is used as a cut off is **40 on 1 method** and **45** on another method within our laboratory.' | Shetland NHS Board |

\*It is not clear if this is a typo error and should read as 37 or is correct

Table 1.1 shows the Cepheid Genexpert test is used by seven trusts which gave five different answers to the threshold - responses included a Ct of 34, 35, 41, 45 and ‘users aren’t provided with the Ct.’ While tests can provide results with high viral load specimens before the full 45 PCR cycles have been completed it is not clear why there is such a disparity in the threshold between trusts for what seems to be the same test.

##### 3.2.2 Cycle threshold is not available

Six responses stated there is no ‘static’, ‘specific’ or ‘standard’ cycle threshold’.’ <u>HS Western Isles</u> reported ‘the exact cycle length is not disclosed in the method literature and is not user-definable.’ See Table 3.2.2.

**Table 3.2.2.**
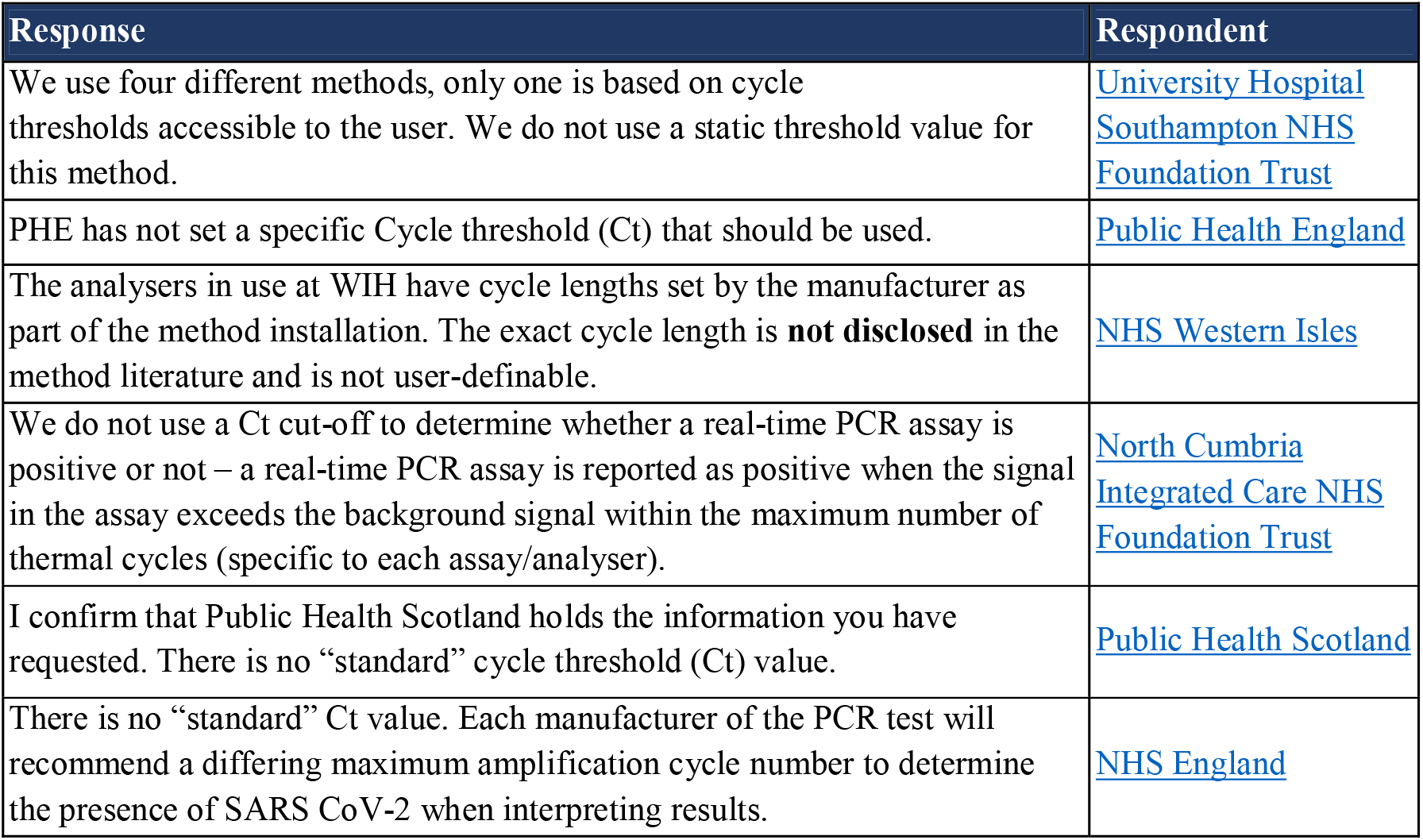
Ct not available.

| Response | Respondent |
| --- | --- |
| We use four different methods, only one is based on cycle thresholds accessible to the user. We do not use a static threshold value for this method. | University Hospital Southampton NHS Foundation Trust |
| PHE has not set a specific Cycle threshold (Ct) that should be used. | Public Health England |
| The analysers in use at WIH have cycle lengths set by the manufacturer as part of the method installation. The exact cycle length is **not disclosed** in the method literature and is not user-definable. | NHS Western Isles |
| We do not use a Ct cut-off to determine whether a real-time PCR assay is positive or not – a real-time PCR assay is reported as positive when the signal in the assay exceeds the background signal within the maximum number of thermal cycles (specific to each assay/analyser). | North Cumbria Integrated Care NHS Foundation Trust |
| I confirm that Public Health Scotland holds the information you have requested. There is no “standard” cycle threshold (Ct) value. | Public Health Scotland |
| There is no “standard” Ct value. Each manufacturer of the PCR test will recommend a differing maximum amplification cycle number to determine the presence of SARS CoV-2 when interpreting results. | NHS England |

One further <u>Trust responded</u> ‘there is no such thing as a cycle threshold.’ However, The Trust did run the assay for 40 cycles and cited this is standard for many PCR assays.

<u>DHSC</u> reports: <u>“some commercial RT-PCR techniques are closed ‘black box’ systems whereby the operator cannot observe the reaction in real-time and the result is interpreted by software into a qualitative non-interrogatable positive or negative result”</u>.

##### 3.2.3 Correlation of cycle threshold with clinical reasoning

We found only one response that correlated the Ct with the clinical picture. (See Table 3.2.3.) Despite the <u>PHE statement</u> that *‘Interpreting single positive Ct values for staging infectious course, prognosis, infectivity or as an indicator of recovery must be done with context about the clinical history*.*’*

**Table 3.2.3.**
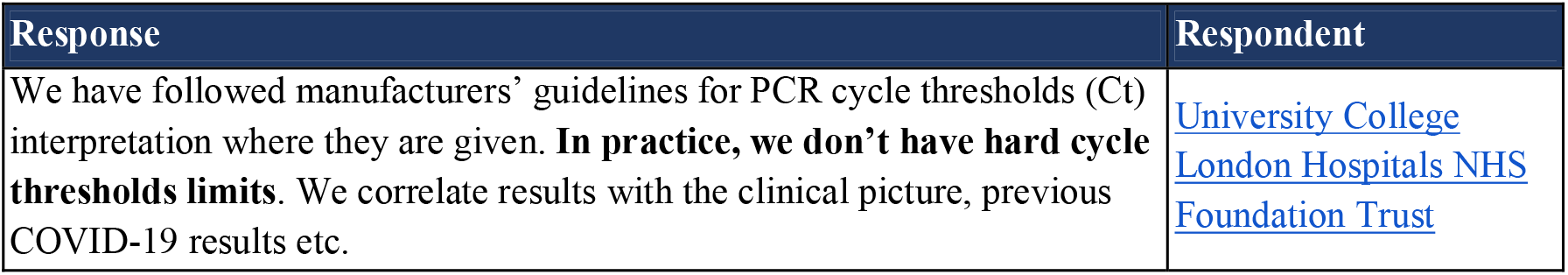
Correlation of Ct with clinical reasoning.

| Response | Respondent |
| --- | --- |
| We have followed manufacturers’ guidelines for PCR cycle thresholds (Ct) interpretation where they are given. **In practice, we don’t have hard cycle thresholds limits.** We correlate results with the clinical picture, previous COVID-19 results etc. | University College London Hospitals NHS Foundation Trust |

DHSC <u>FOI response</u> cites the PHE documents on <u>Understanding cycle threshold (Ct) in SARS-CoV-2 RT-PCR</u> in response to whether ‘the DHSC make/acknowledge any distinction between PCR positives with very high CT cut-offs, and very low CT cut-offs.’

The PHE report considers Ct have a role in identifying infectious individuals that require clinical context:

> *“Low Ct values (high viral load) more likely indicate acute disease and high infectivity*.
>
> *High Ct values (low viral load) can be attributed to several clinical scenarios whereby the risk of infectivity may be reduced but interpretation requires clinical context*.*’*

Public Health Scotland also <u>responded</u> that ‘where available, Ct values are provided to clinicians on a case by case basis”.

### 4. PCR Test Accuracy

#### 4.1 Can you approximate the false positive and false negative rates for your PCR test? Does this vary with the Ct?

PHE responded to an <u>FOI</u> on the known error/accuracy rate with the following quotes:

- ‘RT-PCR tests are highly sensitive and highly specific but can show, as every diagnostic test, low rates of false-negative and false-positive results.’
- ‘These false-negative and false-positive results cannot be entirely eliminated.’
- ‘A test result is called ‘false positive’ when it gives a positive result for a person who is not infected with the infectious agent the test is designed for. ‘
- ‘RT-PCR tests show over 95% sensitivity and specificity. This means that under laboratory conditions, these RT-PCR tests should never show more than 5% false positives or 5% false negatives.’

The false-positive rate (FPR) is the proportion of all those without the disease that yield a positive test outcome. The FPR, therefore, varies with the prevalence of the condition and increases when the prevalence of the condition is low.

Public Health Wales acknowledge this issue in one of their <u>FOI responses</u>:

> ‘*The false positive and false negative rates are determined by the sensitivity and specificity of the test, but also the prevalence of the condition within the test population that is being tested for*.*’*

Public Health Wales acknowledges this issue in one of their <u>FOI responses</u>: ‘The false positive and false negative rates are determined by the sensitivity and specificity of the test, but also the prevalence of the condition within the test population that is being tested for.’

See: <u>Interpreting a covid-19 test result</u>.

In a <u>further response</u>, PHE states that it ‘is very aware of the risk of false positives and that it is working with all Pillar 1 and Pillar 2 labs to put in place measures to reduce that risk including ensuring that appropriate Ct values are being used and that where appropriate samples are re-tested to ensure results are as accurate as possible.

However, we found no reference to what these measures were and to the ‘appropriate CT values’ that can be used to reduce false positives.

One response to a question over whether the trust identified the false positive rate to be of the PCR test to date? The <u>FOI response</u> indicated ‘the diagnostic specificity of the PCR is 100%.’

PHE <u>considers</u> ‘*no diagnostic test is 100% accurate, false positives and false negatives can occur depending on a number of factors not directly related to the test performance*.’ The National technical validation process for manufacturers of SARS-CoV-2 (COVID-19) tests reports eight validated tests with 100% specificity and one test: the Thermo Fisher Taqpath PCR reports 100% (95% CI, 98 to 100%) sensitivity and specificity of 100% (95% CI, 99 to 100%)

Such perfect test results reflect validations based on technical accuracy and not their diagnostic accuracy. Technical accuracy refers to the ability to produce usable information under standardised conditions, whereas diagnostic accuracy depends on the tests intended use in clinical practice.

## Discussion

### Summary for Question 1: How many PCR tests kits are being used in the UK?

Public Health England validation, used early in the pandemic, seemingly, ceased in late 2020. In October 2021, The UK Health Security Agency (UKHSA) was launched as the nation’s new public health body focused on health protection and security.

It is not clear how many test kits there are in use - for most of 2021, PHE reported there were 80 different platforms in use, which increased to 85 in September. DHSC has approved <u>16 tests</u> based on r<u>egulations</u> set out that those supplying COVID-19 tests must apply to DHSC for approval. At the latter end of 2021, a <u>third of all tests</u> in England were analysed in Lighthouse laboratories. However, analysis of European CE marked test kits reveals there are many more tests (over <u>141</u>, and possibly <u>over 400</u>), that might be available for use.

However, there is no central holder of the list of tests, their validation, or their accuracy. The responsibility seemingly lies with the individual laboratories and trusts to oversee which tests they use - only a few responded by naming the tests in use. Therefore, it is currently unclear how many different test kits are in use in the UK and who is responsible for the oversight of which tests are being used in what settings.

### Summary for Question 2: Is there statutory reporting of Ct values with the results of the PCR test?

There is no statutory duty to report Ct values and which tests are being used by which labs to Public Health England or any other body.

Seemingly, there is also no duty by individual laboratories to report what is meant by a “positive,” and all tests may perform differently (especially across laboratories). Not all tests provide a quantitative result: ‘Black box’ systems, for example, are commercial tests that interpret the results using software into a qualitative non-interrogatable positive or a negative result and therefore do not provide Ct results.^2^

The recording of cycle thresholds alongside PCR results is left to the responsibility of the individual hospital trusts. National agencies do not report having a central repository of who does what, and they likely differ substantially in the tests used and how they are reported.

### Summary for Question 3: Do you know what the Ct values are? Do you have a Ct threshold for positivity?

The Department of Health <u>does not hold the information requested</u> but understands that there may be up to 40 cycles in ‘positive’ cases.

<u>Public Health England</u> response contained the standard lamentation on multiple kits used with manufacturer specifications which precludes giving an answer.

Elsewhere, respondents reported variability of cut-offs from 30 to 45 or, in some cases, were invited to ask the manufacturer.

<u>DHSC reports</u> that ‘*Ct values cannot be directly compared between assays of different types due to variation in the sensitivity (limit of detection), the chemistry of reagents, gene targets, cycle parameters, analytical interpretive methods, sample preparation and extraction techniques*.*’*

<u>PHE</u> further reports that ‘every PHE laboratory has in place a strategy for the confirmation of positive results when the screening result was above the threshold identified in the screening assays.’ One Trust provided information on confirmation testing where the sample Ct threshold is >38 on Cepheid Genexpert and >34 Neumodx analysers (see <u>Blackpool Teaching Hospitals NHS Foundation Trust</u>).

Given the limited responses, it is unclear what <u>Non-PHE laboratories</u> do above the threshold limits, as PHE report they do ‘not hold this information for non-PHE laboratories’. It is also unclear what private providers do to confirm results where a positive is above the threshold specified by the manufacturer. There will be considerable variation in the approaches, and some may not undertake confirmatory testing given the time and resources required. It is also unclear to us what tests are used in the nine PHE laboratories and what their performance is.

There also seems to be confusion on the rationale for PCR testing as one response states that “<u>PCR tests are not for viral antigens</u>.”

#### Summary for Question 4: Can you approximate the false positive and false negative rates for your PCR test? Does this vary with the Ct?

We found no information to determine the relationship of test accuracy to how the test would be used in the real world of clinical practice. The false-positive and negative rates are related to the prevalence of the disease, but we found no information answering this question.

There is insufficient information to answer questions relating to the use of the test in the community. For example, results can vary significantly with self-swabbing compared with swabbing by a health professional.

The ONS infection survey reports:^3^

- ‘studies suggest that sensitivity may be somewhere between 85% and 98%.’
- ‘We know the specificity of our test must be very close to 100% as the low number of positive tests in our study over the summer of 2020 means that specificity would be very high even if all positives were false.’

The determination of false positive and false negative rates is currently limited by the lack of comparison with a gold standard. Usually, this should be viral cultures (i.e., growing the SARS-CoV-2 in a safe laboratory from the specimens) and correlating this with the measure of viral burden. However, none of the respondents, when asked, had done this.

The <u>Community prevalence of SARS-CoV-2 in England: Results from the ONS Coronavirus Infection Survey Pilot</u> make this point in their limitations section: ‘ in the absence of a true gold standard, we do not know the test sensitivity and specificity, making it difficult to assess what the true prevalence is.’

## Implications

The answers to the public FOI questions show PHE and DHSC do not know what tests are available; they “validated” only a small number of tests, and each laboratory, outside a small number, are doing their own thing and reporting positive/negative results with no apparent regard to the likelihood of infectiousness.

Instead of running a programme of control, regulation, and validation (governance) of PCR use from the start, the Government allowed the proliferation of PCR tests without a clear understanding of their role and limits.

It is a fact that there is no universal Ct value or cut-off which predicts infectivity. As the major source of the FOI requests, it is essential to understand why this is such a vital issue in the pandemic.

Although a universal cut-off value does not exist, it is unlikely that those “positives” with values over 30 were infectious. The answers indicate that up to a third of those tested were a danger to no one and were presumably isolated for no reason.

Each laboratory and each testing run may give different Ct values from testing the same specimen. However, this is precisely why an uncoordinated system of testing is likely to mislead the public and decision-makers. A positive PCR on its own is not predictive of contagiousness, unless it is integrated with the clinical decision-making pathway of the individual being tested and reports an estimate of viral load as an indication of the likelihood of being infectious.

## Recommendations

- Mandatory control and validation of tests should be introduced.
- In parallel UKHSA should be running a validation programme similar to the one described by <u>Vierbaum et al.</u>
- As with all tests, PCR should never be interpreted outside a clinical context, i.e., a past, recent medical and drug history and careful assessment of exposure to the agent.
- One-off Validation should include calibration of the test against its capacity to detect infectious samples. This should preferentially entail comparison with viral culture.
- No Unvalidated tests should be used.
- A central repository of tests in use and their validation evidence should be published
- Standard reporting on PCR based tests should include an estimated measure of viral burden.
- The reporting of cycle thresholds (or other measures of viral burden) with test positivity should be mandated.
- Any adding or comparison of cases should only include those for whom a reasonable expectation of comparability exists.
- Validation of PCR tests in the real world of clinical practice.
- For the public, we recommend focussing FOI requests to meet section 12 limits (see below).

Significant numbers of FOIs were denied due to the volume of work exceeding the section 12 limits. Section 12 of the FOI Act permits refusals where the cost of compliance exceeds appropriate limits, which the central government sets at £600. This represents the estimated cost of one person spending 24 working hours determining whether the department holds the information, locating, retrieving and extracting the information. See <u>summary description of sections of the FOI Act for refusals</u>

## Conclusion

Access to a response from public health authorities shows there is a lack of knowledge of how many tests are in use, the thresholds used in practice, and a lack of means for identifying contagious individuals. The current system requires significant changes to ensure it offers accurate diagnostic data to enable effective clinical management of SARS-CoV-2.

## Data Availability

All data included in the review are publicly available via Google Docs.

https://docs.google.com/spreadsheets/d/1h2gmiN3kQaFGRy1QMQ8GL6va70UEQ4Fgf2l1PLmSpUc/edit?usp=sharing

